# Prevalence and acceptance of glove wearing practice among general population when visiting high risk are during local COVID-19 outbreak

**DOI:** 10.1101/2020.05.30.20117564

**Authors:** Gobi Hariyanayagam Gunasekaran, Sera Selvanthansundram Gunasekaran, Shargunan Selvanthan Gunasekaran, Fouzia Hanim Bt Abdul Halim, Nur Syafina Insyirah Binti Zaimi, Nor Amirah Binti Abdul Halim

**Affiliations:** Oncology Pharmacy, Hospital Seri Manjung, 32040 Seri Manjung, Perak, Malaysia.; Medical Officer, Hospital Seri Manjung, 32040 Seri Manjung, Perak, Malaysia.; Dental Officer, Manjung District Dental Clinic, 32000 Sitiawan, Perak, Malaysia.

**Keywords:** Covid-19, personal protective equipment, glove, PPE, Malaysia

## Abstract

**Background:** Healthcare authorities have generally advised against wearing glove by the general population. However, the use of gloves has become a common sight in public places raising the question of the necessity of glove wearing practice by the general population

**Objective:** This study aims to investigate the prevalence and types of glove used as well as the acceptance of the glove practice by individuals visiting the high-risk area during Covid-19 pandemic.

**Setting:** This prospective observational study was conducted among individuals visiting a wet market and district specialist hospital During Covid-19 pandemic. The required data was recorded based on observation by trained data collectors who were stationed at the strategic entry point.

**Methods:** Individuals entering through dedicated entry point were observed for the type, category and practice of wearing personal protective equipment. Inclusion criteria for this study were any individuals entering the facilities from entry points without respiratory symptoms. Exclusion criteria for this study were individuals less than 2 years old, visiting the emergency department, facility staff, individuals who are suspected of multiple entry and individuals who are exiting the treatment facility entrance. Patients were categorized into two groups of acceptable and unacceptable glove practice. The Pearson chi-square was used to test for differences in investigated variables in the univariate setting.

**Main outcome measure:** Prevalence, acceptance of glove wearing practice.

**Results:** A total of 75 individuals (2.3%) compromising of 45 (60.0%) individuals from hospitals and 30 (40.0%) individuals from wet markets were seen wearing glove amongst 3322 individuals observed during the data collection period. A higher proportion of individuals visiting wet market (30.0%) were observed with unacceptable glove practice compared to individuals visiting the hospital (8.9%), χ^2^ (1) = 5.60, p = .018. Similarly, a Higher proportion of glove use among non-Malay (53.3%) compared to Malay (46.7%) was observed in hospital compared to a higher proportion of glove use among Malay compared to non-Malay (16.7%) visiting wet market, χ^2^ (1) = 10.20, p = .001. As for glove use, we found that male were using more medical-grade glove (78.8%) compared to non-medical grade glove (21.2%) while an equal amount of medical (50.0%) and non-medical grade glove (50.0%) was used among female, χ^2^ (1) = 6.546, p = .011. Besides, we found that higher proportion of individual using medical-grade glove was using medical grade facemask (68.3%) which was similar to the proportion of individuals using non-medical glove was using non-medical facemask (66.7%), χ^2^ (1) = 5.25, p = .022.

**Conclusion:** We present the prevalence and characteristics of glove wearing practice in high-risk location during the current COVID-19 outbreak in Malaysia. Facing a worldwide public health emergency with limited effective clinical treatment, the role of glove-wearing in mitigating COVID-19 transmission is questionable. If needed, the compliance to proper glove-wearing could be improved through targeted public health education

## Introduction

COVID-19 coronavirus (also known as 2019-nCoV or SARS-CoV-2) has spread dramatically worldwide overwhelming health-care systems[1, 2]. While no known effective treatment is available, use of personal protective equipment has been advocated by health care authorities to reduce the risk of spreading the virus by human-to-human transmission[3–5] or through contaminated surfaces [6].

Early reports have concluded that Human-to-human transmissions of SARS-CoV-2 were facilitated through respiratory droplets from coughing and sneezing[7, 8]. Subsequently, health care authorities have promoted the use of facemask among the general population as paramount PPE to contain this epidemic [7–13]. Recent studies on SARS-CoV-2 have suggested environment surface as a potential medium of transmission as the virus was detectable on a variety of surfaces from hours to days[14, 15]. A recent study by Ong et al. evaluated multiple air and surface samples from COVID-19 isolation wards reported positive samples were found on common environmental surfaces such as the table, bed rail, locker, chair, light switches, door, window, and surfaces in the toilet including the toilet bowl, sink, and door handle[6]. Besides, Otter and his colleagues found that SARS-CoV-2 and other coronaviruses can survive on environmental surfaces up to 6 days[16]. This finding implies that a person has chances of being infected after touching the objects contaminated by SARS-CoV-2 virus although the actual risk is unknown.

Reports of a surface contact as a possible route of transmission have raised a question on the need to use gloves as PPE among general population. The risk of hand hygiene as a potential risk factor for transmission have been highlighted by Kwok YL and team who have reported that individuals commonly touch their faces 23 times per hour and 44% of those touches involved contact with mucous membranes[17]. Lately, news of shops requiring customers to wear both facemask and glove was reported [18, 19]. Concurrently, the anticipated increase of glove use by general population have been lead to an increased global production of glove[20]

Generally, health care authorities have not advised the general population to use a glove in public areas. The World Health Organisation does not recommend the use of gloves[21] while healthcare service executive of Ireland advises against wearing disposable gloves [22]. Centre for disease prevention and control have only suggested the use of gloves when for surface disinfection in contact with suspected or confirmed COVID-19 patient[23].

Although the use of glove is generally not recommended, the use of disposable, single-use plastic or latex gloves has become a common sight in public places, particularly in high-risk areas such as hospital and markets. However, the use of glove by the general population is still unknown. Thus the preliminary result of glove use of this research could be used to improve

## Aim of the study

This study aims to investigate the prevalence and types of glove used as well as the acceptance of the glove practice by individuals visiting high risk area during Covid-19 pandemic.

## Ethics Approval

The ethical approval to conduct the study was obtained from the Medical Ethical Review Committee [MERC KKM. NIHSEC. P20-902(6)] and [MERC KKM. NIHSEC. P20-1002(6)] Ministry of Health, Malaysia.

## Methods

### Study setting

This prospective observational study was conducted among individuals visiting a wet market and district specialist hospital in Sitiawan, Perak, Malaysia. During Covid-19 pandemic, entrance to the facility was limited via the dedicated entry point while all other peripheral entrance was closed to control the movement of individual entering and exiting the facilities. The required data was recorded based on observation by trained data collectors who were stationed at strategic entry point.

### Inclusion and Exclusion

Inclusion criteria for this study were any individuals entering the facilities from entry points without respiratory symptoms. Exclusion criteria for this study were individuals less than 2 years old, visiting emergency department, facility staff, individuals which are suspected of multiple entry and individuals who are exiting the treatment facility entrance. strategic management for public health.

### Data Variables

Individual data were collected by visually observing the type of the glove and evaluating the acceptance of glove use among visitors entering into the facility. Demographic data such as patient’s gender, age group and ethnicity while glove data such as category and acceptance of glove practice was recorded. Gender was categorised as either male or female while patients ethnicity was categorised into Malay or Non-Malay to reflect population distribution (24). The visitor’s age group was recorded as either as children, adult or elderly which was done based on subject’s facial and physical feature (25). The age group was further categorised to low-risk age (children and adult) or high-risk age (elderly) group (26–28). Glove usage was classifies as either “Yes” when the any type of glove is used or as “No” when the glove is absent. The category of glove used was described according to their class; latex, nitrile, cloth or plastic. The glove was further categorized as medical-grade (latex and nitrile glove) or non-medical grade (cloth and plastic).The acceptance level of garbing practice for glove was recorded as acceptable (correct usage) practice or unacceptable (incorrect usage) practice. The reason for unacceptable practice was further describes as well.

### Statistical analysis

All demographic and categorical variables were presented as number (n) and percentage (%). Pearson’s chi-squared test was used to determine the statistically significant difference between the demographic characteristic between age group and acceptance level of glove garbing practice. Simple logistic regression was used to screen independent variable. Variables with p value < 0.25 was included in the multivariate analysis. Binomial logistic regression test was applied to determine the contributing factor to unacceptable glove practice. Correlation matrix was checked for interaction between the variables. The Hosmer and Lemeshow test, and Classification table was used to evaluate model of good fit. The final model was presented with 95% confidence interval (CI) and its corresponding p-value. For all test Two-tailed p-value < 0.05 was considered as statistically significant. All statistical analyses were performed using SPSS for Windows version 22.0 (SPSS Inc., Chicago, Illinois, USA).

### Result

At total of 75 individuals (2.3%) compromising of 45 (60.0%) individuals from hospitals and 30 (40.0%) individuals from wet markets were seen wearing glove amongst 3322 individuals observed during the data collection period. More female (56.0%) was observed wearing glove compared to male (44.0%) individuals. As for ethnicity higher proportion of Malay ethnic (61.3%) was observed wearing glove compared to non-Malay ethnic (38.7%). Majority of the individuals were from low-risk age group (82.7%) which comprised of children (6.7%) and adult (76.0%) category while the remaining in high-risk age group (17.3%) were from elderly category. As for glove use, higher proportion of them were using medical grade glove (62.7%) while the remaining was using non-medical grade glove (37.3%). This study group also noticed that all 75 individuals included in this study was wearing facemask. Among them, 63 individuals (84.0%) was wearing medical grade face such as surgical grade facemask (81.3%) and respirator type facemask (2.7%) while the remaining 12 individual (16.0%) was wearing non-medical grade facemask such as cloth facemask (10.7%) and paper facemask (5.3%).

**Table 1.** Demographic characteristic, glove type and usage among general population during visit to high-risk areas

| Description | Frequency (n=75) | Percentage (%) |
| --- | --- | --- |
| **Facility** |  |  |
| Hospital | 45 | 60.0 |
| Wet market | 30 | 40.0 |
| **Gender** |  |  |
| Male | 33 | 44.0 |
| Female | 42 | 56.0 |
| **Ethnic** |  |  |
| **Malay** |  |  |
| Malay | 46 | 61.3 |
| **Non-Malay** |  |  |
| Chinese | 17 | 22.7 |
| Indian | 12 | 16.0 |
| **Age Group** |  |  |
| **Low-risk Age group** |  |  |
| Children | 5 | 6.7 |
| Adult | 57 | 76.0 |
| **High-risk Age group** |  |  |
| Elderly | 13 | 17.3 |
| **Category of Glove** |  |  |
| **Medical Grade** |  |  |
| Latex | 19 | 25.3 |
| Nitrile | 28 | 37.3 |
| **Non-Medical Grade** |  |  |
| Cloth | 4 | 5.3 |
| Plastic | 24 | 32.0 |

Table 2 describes the distribution of variables according to acceptance of glove practice and demographic variables. Among 75 individuals using glove, 62 individuals (82.7%) had acceptable glove practice while the remaining 13 individuals (17.3%) had unacceptable glove practice.

**Table 2.** Demographic characteristic between acceptance of glove use (n = 75) Description Acceptable (n = 62)

| **Table 2** Demographic characteristic between acceptance of glove use (n=75) |  |  |  |  |  |
| --- | --- | --- | --- | --- | --- |
| Description | Acceptable (n=62) |  | Unacceptable (n=13) |  | p-value |
|  | Frequency | Percentage | Frequency | Percentage |  |
| **Facility** |  |  |  |  | .018 |
| Hospital | 41 | 91.1 | 4 | 8.9 |  |
| Wet market | 21 | 70.0 | 9 | 30.0 |  |
| **Gender** |  |  |  |  | .863 |
| Male | 27 | 81.8 | 6 | 18.2 |  |
| Female | 35 | 83.3 | 7 | 16.7 |  |
| **Ethnic** |  |  |  |  | .520 |
| Malay | 37 | 80.4 | 9 | 19.6 |  |
| Non-Malay | 25 | 86.2 | 4 | 13.8 |  |
| **Age group** |  |  |  |  | .838 |
| Low-risk | 51 | 82.3 | 11 | 17.7 |  |
| High-Risk | 11 | 84.6 | 2 | 15.4 |  |
| **Category of glove** |  |  |  |  | .590 |
|  | 38 | 80.9 | 9 | 19.1 |  |
| Medical Grade | 24 | 85.7 | 4 | 14.3 |  |
| Non-Medical Grade |  |  |  |  |  |

A significant difference in distribution between acceptance of glove practice and demographic variable of facility and type of glove was observed. Higher proportion of individuals visiting wet market (30.0%) were observed with unacceptable glove practice compared to individuals visiting hospital (8.9%), χ^2^ (1) = 5.60, p = .018. Similarly, statically significant difference between ethnic and visit to study location among individuals wearing glove was observed as well. Higher proportion of glove use among non-Malay (53.3%) compared to Malay (46.7%) were observed in hospital compared to higher proportion of glove use among Malay compared to non-Malay (16.7%) visiting wet market, χ^2^ (1) = 10.20, p = .001. As for glove use, we found that male were using more medical grade glove (78.8%) compared to non-medical grade glove (21.2%) while equal amount of medical (50.0%) and non-medical grade glove (50.0%) was used among female, χ^2^ (1) = 6.546, p = .011. in addition we found that higher proportion of individual using medical grade glove was using medical grade facemask (68.3%) which was similar to proportion of individuals using non-medical glove was using non-medical facemask (66.7%), χ^2^ (1) = 5.25, p = .022.

Among the 13 subjects with unacceptable glove practice, all of them were wearing glove only on one side of hand. We also observed difference in facemask practice among our study population, Among 62 patients with acceptable glove practice 3 of them had unacceptable facemask practice (either mouth or nose was uncovered). On the contrary, among 13 individuals with unacceptable glove practice, all of them had acceptable facemask practice.

## Discussion

Our study describes the types and acceptance of glove wearing practice during local COVID-19 outbreak. The prevalence of glove wearing was 2.3% of the observed study population. We were not able to compare the prevalence of our study population as there are no other studies which have reported the glove use among the non-healthcare population.

Although the prevalence of glove use among our study population was small, undeniably the uptake of the use of glove among the general population has increased due to COVID-19 outbreak. There have been reports of substantial increase in the use of PPE both in community and healthcare settings once the local outbreak begins [24–26]. Public perception on the necessity to wear PPE was fuelled by media coverage of graphic pictures of civilian, authorities and health care personnel wearing extensive personal protective equipment (PPE) During the early stage of the outbreak [27]. reports of SARS-CoV-2 virus originating from wet market and reports of SARS-CoV-2 virus detected on surfaces in patient rooms [28] might have encouraged glove wearing practice.

As SARS-CoV-2 virus transmitted predominantly by droplets, local health authorities recommendation of facemask use in the public location was universally headed, evidently as facemask coverage of 96.9%-99.7% among local population was reported [29, 30]. Similarly, the low proportion of glove use could be contributed by the recommendation against use of glove by local health authorities [31] which is similar to stand taken by other public healthcare agencies such as World Health Organisation [21], healthcare service executive of Ireland [22] and Centre for disease prevention and control (CDC, USA)[23].

Although the use of glove among general population is negligible, unacceptable glove practice among individuals visiting wet market (30.0%) and hospital (8.9%) raises the question on the necessity of glove use in community setting. To date, there are no clinical evidence accepting or refuting the benefit of glove wearing among public in relation to Covid-19 pandemic. However, anecdotal evidence does not support the benefit of glove use outside medical setting. In addition, various doctors have cautioned that use of glove might provide false sense of security and divert attention from the importance of washing hands[18, 31, 32]. This is in concord with little evidence that wearing gloves in public offers protection from contracting the virus. While intentions are good, the indiscriminate wearing of gloves of general public is largely ineffective and may lead to more harm than good. If needed, the compliance to proper glove wearing could be improved through targeted public health education[33, 34].

We found that higher proportion of male was using medical grade glove (78.8%) compared to non-medical grade glove (21.2%).The higher proportion of medical grade glove use among male were likely due to the widespread news of higher risk of mortality among male gender[35–37], with reports of Males are 1.85 times (OR: 1.85, 95% CI 1.60–2.13) more likely to die from COVID-19 infection [38]. This high risk of death among male gender might have promoted extra protection seeking behaviour among this group of patients. This is evident as not only we have observed universal facemask practice among our study population, we also have observed higher proportion (68.3%) of individual using medical grade glove was using medical grade facemask as well. Although the use of glove is in individual dependent, the mental wellbeing contributing to glove usage should not be neglected[39]. For example, Lin et al. correlated all-time high search for “face mask” in Google could be a sign of anxiety appearing in the society[40]. Szczesniak et al. and teams findings imply that in addition of protection against the COVID-19, the use of facemask also increase the level of perceived self-protection and of social solidarity which, thereby improve mental health wellbeing[41] similar to Correspondence of Shannon L et al have described similar sense increased morale within her department and promoted buy-in to the use of PPE [42]. Though glove use was not recommended, public might still chose to wear glove to boost morale or sense of protection.

Based on the above evidence, hand hygiene, together with appropriate personal protective equipment, is of utmost importance to break the cycle of touching contaminated environmental surfaces and subsequent inoculation of the virus through touching mucous membranes, thus reducing the risk of transmission of COVID-19.

The finding our study might be biased due to the study location. Wet markets and hospitals are generally perceived as a high risk location for SARS-CoV-2 virus transmission. Thus, visitors to such location might have taken extra precaution such as wearing glove which might not be practiced in other circumstance. Besides that, the availability and price of glove on market could have greatly influenced subject’s preference.

## Conclusion

We present the prevalence and characteristics of glove wearing practice in high risk location during the current COVID-19 outbreak in Malaysia. Facing a worldwide public health emergency with limited effective clinical treatment, the role of glove wearing in mitigating COVID-19 transmission is questionable. If needed, the compliance to proper glove wearing could be improved through targeted public health education

## Data Availability

The data that support the findings of this study are available from the corresponding author, upon reasonable request.

## Notes

### Competing Interest Statement

The authors have declared no competing interest.

### Funding Statement

No funds were obtained for study design, data collection and analysis, the decision to publish, or preparation of the manuscript.

